# Disentangling adiposity-related and non-adiposity-related genetic pathways for type 2 diabetes

**DOI:** 10.64898/2026.06.11.26355425

**Authors:** Chen-Yang Su, Tianyuan Lu

**Affiliations:** Department of Population Health Sciences, School of Medicine and Public Health, University of Wisconsin-Madison, Madison, WI, USA; Department of Biostatistics and Medical Informatics, School of Medicine and Public Health, University of Wisconsin-Madison, Madison, WI, USA; Center for Precision Medicine, University of Wisconsin-Madison, Madison, WI, USA; Center for Genomic Science Innovation, University of Wisconsin-Madison, Madison, WI, USA; Center for Demography of Health and Aging, University of Wisconsin-Madison, Madison, WI, USA

## Abstract

**OBJECTIVE:** To identify circulating proteins associated with type 2 diabetes (T2D) risk through pathways not fully explained by body mass index (BMI), and to assess therapeutic actionability.

**RESEARCH DESIGN AND METHODS:** We applied GWAS-by-subtraction within a genomic structural equation model to European ancestry summary statistics for T2D (74,124 cases, 824,006 controls) and BMI (n = 681,275), partitioning T2D liability into BMI-related and BMI-subtracted components. We then performed proteome-wide Mendelian randomization (MR) using *cis*-protein quantitative trait loci from four plasma proteomics cohorts: ARIC, deCODE, Fenland, and the UK Biobank Pharma Proteomics Project. Prioritized proteins passed sensitivity analyses with alternative MR methods and were supported by colocalization evidence. Tissue-resolution regulatory support was assessed using *cis*-eQTL colocalization across GTEx and pancreatic islet, subcutaneous adipose, and whole-blood resources. Actionability was evaluated using the druggable genome and Open Targets.

**RESULTS:** GWAS-by-subtraction attenuated the genetic correlation between BMI and BMI-subtracted T2D from 0.54 (SE 0.02) to 0.35 (SE 0.02). Proteome-wide MR prioritized 29 proteins for BMI-subtracted T2D. Thirteen showed eQTL colocalization in at least one tissue, implicating liver and intermediary metabolism (GCDH, NOTCH2), pancreatic islet biology (CTRB2, MANBA), adipose and Wnt signaling (RSPO3, GALNT3), and whole blood regulatory signals (PAM, SNUPN). Sixteen proteins were classified within druggable-genome Tiers 1-3, and five had existing Open Targets compounds.

**CONCLUSIONS:** Integrating GWAS-by-subtraction, proteome-wide MR, and colocalization nominated 29 proteins associated with T2D liability not fully explained by BMI. These findings highlight genetically supported targets for follow-up studies of T2D therapies that complement weight-centered approaches.

**Article Highlights:** - **Why did we undertake this study?** T2D is clinically and biologically heterogeneous. Although human genetics can prioritize disease-relevant pathways, T2D genetic associations are strongly influenced by body mass index (BMI), which obscures identification of targets contributing to T2D beyond adiposity.
- **What is the specific question we wanted to answer?** Which circulating proteins are associated with non-BMI-mediated T2D, and how actionable are they as therapeutic targets?
- **What did we find?** GWAS-by-subtraction, proteome-wide Mendelian randomization, and tissue-resolution colocalization prioritized 29 proteins for BMI-subtracted T2D, implicating metabolic, pancreatic-islet, signaling, and glycosylation-related pathways, with many representing pharmacologically tractable targets.
- **What are the implications of our findings?** These results nominate genetically informed protein targets for non-BMI-mediated T2D pathways, supporting follow-up studies of therapies that complement weight-centered approaches.

## Introduction

Type 2 diabetes (T2D) is a major global health challenge, affecting over 500 million individuals worldwide and imposing a substantial clinical and economic burden (1). Despite substantial therapeutic progress, including the widespread use of GLP-1 receptor agonists and SGLT2 inhibitors (2,3), T2D remains clinically and biologically heterogeneous, and important unmet therapeutic needs persist (4).

Human genetic evidence provides an opportunity to prioritize disease-relevant biology and nominate therapeutic targets. Genome-wide association studies (GWAS) have identified hundreds of loci associated with T2D risk, providing important insights into disease biology (5,6). When integrated with molecular trait data, these discoveries can be used in Mendelian randomization (MR), an approach that uses genetic variants associated with an exposure, such as circulating protein abundance, to estimate whether genetically predicted differences in that exposure are associated with disease risk (7). Proteome-wide MR can therefore help prioritize proteins with putative causal effects on T2D and nominate targets with potential pharmacological relevance.

However, MR-facilitated target discovery depends critically on the GWAS used to define the disease outcome. Many T2D-associated loci are also associated with adiposity-related traits, and genetic correlation analyses have demonstrated substantial shared genetic architecture between BMI and T2D (8,9). As a result, proteome-wide MR using conventional T2D GWAS may preferentially identify proteins and pathways whose effects on T2D are mediated by BMI-related biology. Conventional GWAS of T2D aggregate genetic effects that act through BMI-related pathways, such as insulin resistance driven by adiposity, with those that reflect non-BMI-mediated mechanisms, such as pancreatic β-cell dysfunction. This conflation limits the utility of human genetic evidence for discovering targets relevant to patients whose underlying disease mechanisms do not primarily act through BMI, including individuals who develop T2D in the absence of obesity or in settings such as lipodystrophy.

Approaches that explicitly partition shared genetic architecture between correlated traits address this limitation. In particular, GWAS-by-subtraction is a multivariable genomic structural equation modeling approach that decomposes genetic liability for an outcome into components shared with, or residual to, a correlated trait such as BMI (10,11), and may mitigate the risk of collider bias when adjusting for a heritable trait in covariate-adjusted GWAS (12). By separating BMI-related from non-BMI components of T2D genetic liability, this can enable downstream genetic analyses to focus on disease biology not fully captured by adiposity.

Here, we applied GWAS-by-subtraction to partition T2D genetic liability into BMI-related and non-BMI-mediated components. We then combined proteome-wide Mendelian randomization, tissue-specific colocalization, and actionability assessment to identify circulating proteins with putative causal effects on non-BMI-mediated T2D risk. This approach aimed to prioritize pharmacologically tractable protein targets that may inform complementary strategies for T2D prevention and treatment beyond adiposity-centered pathways.

## Research Design and Methods

### Data Sources

For T2D, we used a European ancestry meta-analysis of T2D that was unadjusted for BMI (*n* = 898,130; 74,124 T2D cases and 824,006 controls) (6). For BMI, we used GWAS from the GIANT consortium and UK Biobank meta-analysis, including 681,275 individuals of European ancestry (8). Plasma proteomics were obtained from four European-ancestry cohorts: the Atherosclerosis Risk in Communities (ARIC) study (n = 7,213; 4,657 proteins) (13), deCODE (n = 35,559; 4,719 proteins) (14), Fenland (n = 10,708; 4,775 proteins) (15) which were measured on the SomaScan v4 platform, and the UK Biobank Pharma Proteomics Project (UKB-PPP; n = 34,557; 2,923 proteins) using Olink Explore 3072 (16). *Cis*-eQTL summary statistics were obtained from GTEx v8 for six T2D-relevant tissues (pancreas, liver, skeletal muscle, subcutaneous adipose, visceral adipose, and whole blood) (17), and from three tissue-specific resources: InsPIRE consortium pancreatic islets (n = 420) (18), METSIM subcutaneous adipose (n = 434) (19), and the eQTLGen Phase I whole-blood meta-analysis (up to n = 31,684) (20). These data are shown in **Supplementary Note 1**.

### GWAS-by-subtraction

We applied GWAS-by-subtraction using GenomicSEM v0.0.5 (10,11) which uses linkage disequilibrium score regression (LDSC)-estimated heritabilities and the genetic covariance between T2D and BMI as input to a two-trait genomic structural equation model. Detailed model specification have been previously described (21,22). We partitioned T2D genetic effects into a BMI-related latent component (genetic effects shared with BMI) and a BMI-subtracted component (residual effects after accounting for BMI-related effects). Genome-wide association testing was then performed for each latent component. Genome-wide significance was defined as *P* <5×10^-8^.

### Genetic Correlation Analyses

We used LDSC (23) and the European 1000 Genomes Project reference to estimate heritability, intercepts, and genetic correlations between BMI and the original T2D GWAS, the BMI-related component, and the BMI-subtracted component.

### Proteome-wide Mendelian randomization

Independent *cis*-pQTLs were identified per cohort by LD clumping (*P* <5×10^-8^; *r*^2^ <0.001), restricting to variants within ±500 kb of the transcription start site of the encoding gene. Variants in the major histocompatibility complex were excluded. Two-sample MR was performed using inverse-variance weighting (IVW) for proteins with ≥2 instruments and the Wald ratio otherwise (24). Multiple testing was primarily controlled using a stringent study-wide Bonferroni threshold to define the main prioritized protein set, with within-cohort Benjamini-Hochberg false discovery rate correction reported as an additional complementary analysis. Sensitivity analyses for proteins with ≥3 instruments included weighted-median (25), weighted-mode (26), and MR-Egger estimators (27), MR-Egger intercept tests for directional pleiotropy, Cochran’s Q for heterogeneity, and Steiger filtering for reverse causation. Reporting followed the STROBE-MR guidelines (**Supplementary Note 2**).

### Colocalization

Colocalization between *cis*-pQTL and BMI-subtracted T2D signals was performed within ±500kb using two complementary methods, PWCoCo (28) and SharePro (29), that account for multiple causal variants. We used the 1000 Genomes Project European ancestry cohort as the LD reference. Following our previous work, targets were retained when at least one method returned a posterior probability of a single shared causal variant (PP.H4) ≥ 0.8 (30).

For each prioritized protein, we further evaluated colocalization between the corresponding *cis*-pQTL and *cis*-eQTL signals across the six GTEx tissues and three tissue-specialty resources listed above. eQTL data from InsPIRE, METSIM, and eQTLGen were obtained as raw nominal cis-eQTLs, lifted from hg19 to hg38 using UCSC liftOver, and harmonized; for eQTLGen, β and SE were derived from Z-scores using Zhu et al. (31). For each prioritized protein, we considered eQTL colocalization PP.H4 across the proteomic cohorts in which the protein passed the MR Bonferroni threshold and report the strongest PP.H4 per tissue.

### Functional annotation and network analysis

We also manually annotated each prioritized protein into broad mechanistic modules based on canonical protein function, tissue or cell-type context, GWAS locus biology, and experimental or human genetic evidence from a literature search. To interpret the 29 Bonferroni- and colocalization-passing proteins, we assessed protein-protein interaction enrichment using STRING v12 (32) and pathway over-representation using g:Profiler/gprofiler2 (33) across Gene Ontology, KEGG, and Reactome. STRING analyses retained medium-confidence interactions (combined score ≥0.4). Whole-network PPI enrichment was assessed using STRING’s built-in PPI enrichment test, which compares the observed number of medium-confidence edges among the 29 query proteins to the number expected for a same-size random set of proteins drawn from the human interactome with each query protein’s degree preserved, and reports a one-sided upper-tail P-value. PPI enrichment was considered significant at *P* <0.05. To limit chance over-representation of small overlaps, we required at least three prioritized genes in the query intersection and excluded ontology root terms. For g:Profiler, we used the annotated genome as the enrichment background. Enrichment was defined as g:SCS-adjusted *P* <0.05.

### Actionability assessment

The druggable-genome (34) was used to stratify proteins into Tiers 1 (approved or clinical-stage targets), 2, 3A, and 3B, with unclassified proteins flagged separately. Drug repurposing opportunities were further annotated using the Open Targets Platform (35) for known compound interactions and the most advanced clinical trial phase per target.

### Ethics

This research was conducted using secondary data and has been determined to be exempt from IRB review under the University of Wisconsin-Madison Institutional Review Board policies with a formal IRB waiver. All summary level data are publicly available.

### Data and Resource Availability

Analyses were performed in R v4.5.2 (https://www.r-project.org/). We used GenomicSEM (https://github.com/GenomicSEM/GenomicSEM), UCSC liftOver, PLINK v1.9 (http://pngu.mgh.harvard.edu/purcell/plink/), TwoSampleMR v0.7.5 (https://mrcieu.github.io/TwoSampleMR/), coloc v5.2.3, PWCoCo, SharePro v5.0 (https://github.com/zhwm/SharePro_coloc).

## Results

### GWAS-by-subtraction separates T2D genetic associations from BMI-related effects

We applied GWAS-by-subtraction to partition common variant associations with T2D into components shared with BMI, termed “BMI-subtracted”, and components that remained after accounting for BMI-related genetic effects, termed “BMI-related” (**Figure 1a**). In line with the expected contribution of adiposity to T2D risk, the BMI-related component showed more polygenic genome-wide associations than the BMI-subtracted component (**Figure 1b**). The BMI-subtracted T2D GWAS had an LDSC intercept of 1.01 (SE = 0.03), confirming that the remaining signals were unlikely to be driven by population stratification or other forms of confounding. The BMI-related T2D component similarly had an intercept close to one (1.00, SE = 0.03) (**Supplementary Table 1**). Consistent with the stronger genome-wide signal in the BMI-related component, we detected 38,869 genome-wide significant SNPs (*P* <5×10^-8^) for BMI-related T2D effects compared with 4,871 genome-wide significant SNPs for BMI-subtracted T2D effects (**Supplementary Table 2** and **3**).

**Figure 1.**
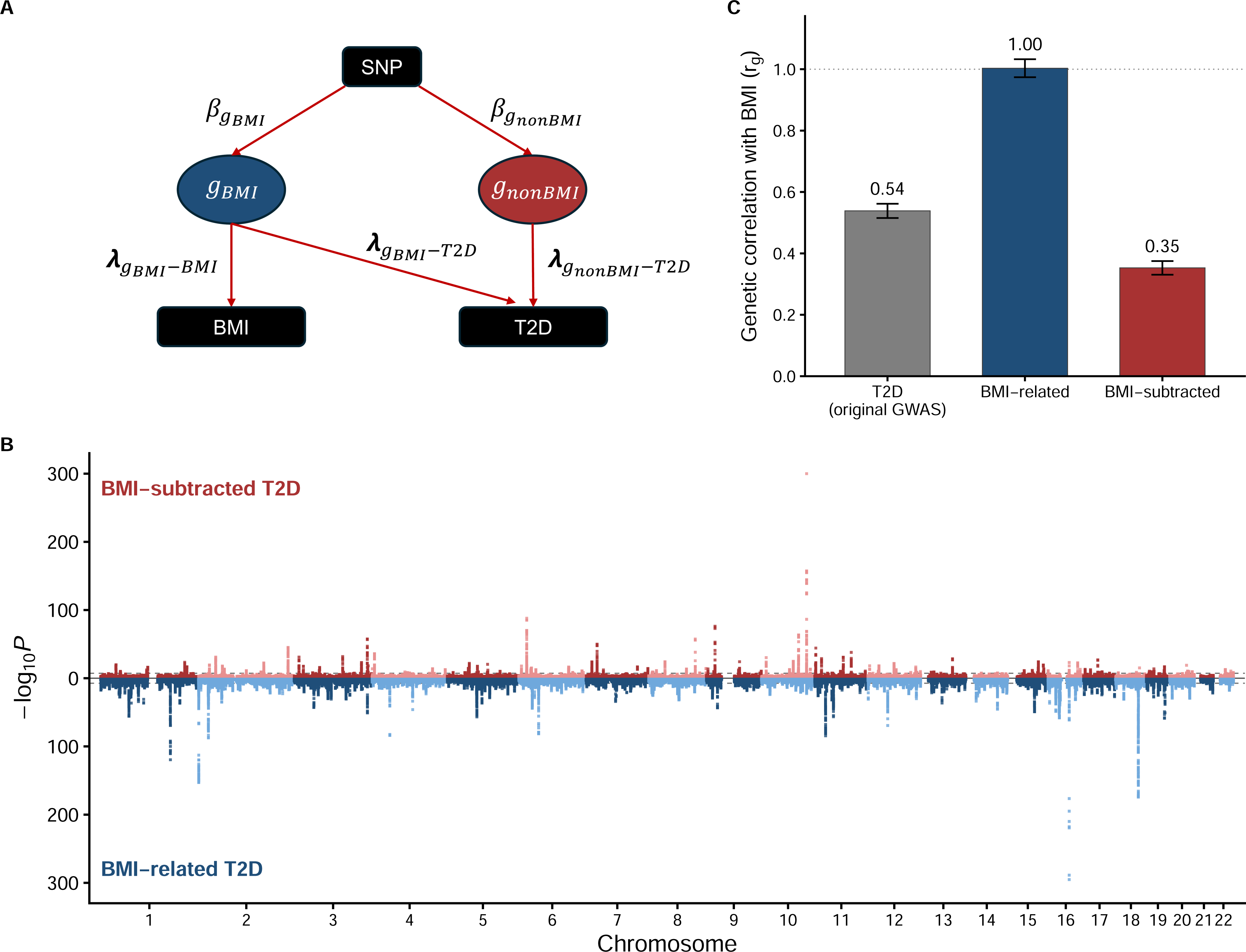
Genetic architecture of BMI-subtracted and BMI-related T2D. (A) Illustration of GWAS-by-subtraction model for BMI-derived effects on type 2 diabetes (T2D). <u>Genetic latent factors</u> *g_BMI_*: BMI genetic latent factors that may include T2D genetic effects *g_nonBMI_*: non-BMI genetic latent factors <u>SNP to genetic latent factor effects</u> *β_gBMI_*: SNP effect on BMI genetic latent factor *β_gnonBMI_*: SNP effect on non-BMI genetic latent factor <u>Genetic latent factor to trait effects</u> *λ_gBMI-BMI_*: effect of BMI genetic latent factors on genetic components of BMI *λ_gBMI-T2D_*: effect of BMI genetic latent factors on genetic components of T2D *λ_gnonBMI-T2D_*: effect of non-BMI genetic latent factors on genetic components of T2D (B) LDSC genetic correlations (*r_g_*) with BMI for the original T2D GWAS, the BMI-related T2D component, and the BMI-subtracted T2D component. Error bars indicate ± 1 SE. Bars show the point estimate, error bars indicate ±1 SE, and the dotted line marks *r_g_*= 1. (C) Miami plot of BMI-subtracted T2D (top) and BMI-related T2D (bottom) summary statistics. Each point represents a SNP plotted by chromosomal position (x-axis) and -log_10_*P*-vaue (y-axis); alternating colors distinguish chromosomes; the dashed line indicates the genome-wide significance threshold (*P* = 5 × 10^-8^).

Among the variants reaching genome-wide significance in the BMI-subtracted T2D GWAS, many associations were weaker than those in the BMI-related component, indicating that part of the T2D signal is captured by BMI-associated pathways. However, a distinct set of variants retained strong residual association after subtraction, supporting the presence of T2D genetic risk that is not fully explained by BMI. The strongest BMI-subtracted signal was observed at the *TCF7L2* locus on chromosome 10 (rs7901695, effect allele T, MAF = 0.34; OR = 0.85, 95% CI 0.85-0.86; *P* <1×10^-300^) which is the largest effect common-variant locus for T2D and a regulator of pancreatic β-cell function. The strongest BMI-related signal was observed at *FTO*, a canonical adiposity-driving locus, on chromosome 16 (rs7206790, effect allele C, MAF = 0.50; OR = 0.98, 95% CI 0.98-0.98; *P*=5.96×10^-296^). More broadly, many T2D loci showed appreciable attenuation upon removal of BMI-related effects, indicating that a substantial portion of the genome-wide signal at these loci is captured by adiposity-linked pathways. By contrast, a meaningful subset of loci retained significant associations after subtraction, suggesting contributions to T2D susceptibility not mediated through BMI.

To validate that GWAS-by-subtraction partitioned BMI-related effects as intended, we used LDSC to estimate the genetic correlation between BMI and (i) the original T2D GWAS, (ii) the BMI-related T2D component, and (iii) the BMI-subtracted T2D component (**Figure 1c**). Prior to subtraction, BMI and T2D were strongly correlated (*r_g_* = 0.54, SE = 0.023), consistent with the well-established epidemiological link between adiposity and T2D and confirming that a sizeable fraction of common variant T2D risk is shared with BMI. As expected, the BMI-related T2D component was perfectly correlated with BMI (*r_g_* = 1.00, SE = 0.029), confirming that this latent factor captures the BMI-mediated effects. Crucially, after subtracting BMI effects, the genetic correlation between BMI and BMI-subtracted T2D genetic effects was attenuated (*r_g_* = 0.35, SE = 0.022), suggesting potential shared upstream regulators of metabolic health. Together, these findings support GWAS-by-subtraction as a useful approach for distinguishing BMI-related T2D genetic effects from residual T2D associations that may reflect non-BMI-mediated pathways.

### Proteome-wide MR and Colocalization Prioritize Candidate Proteins for BMI-subtracted T2D

To identify circulating proteins associated with T2D liability not captured by BMI, we performed proteome-wide MR using *cis*-pQTL from ARIC, deCODE, Fenland, and UKB-PPP against BMI-subtracted T2D (**Supplementary Table 4-7**). Instrument strength was high across cohorts, with cohort-level median F-statistics ranging from 174 to 328. The minimum F-statistic was 30 across all cohorts, indicating low susceptibility to weak-instrument bias (**Supplementary Table 8** and **9**).

Across the four proteomic cohorts, we performed 4,991 tests, comprising 1,166 tests in ARIC, 1,202 in deCODE, 1,214 in Fenland, and 1,409 in UKB-PPP. We therefore applied a study-wide Bonferroni threshold of *P* < 0.05/4,991, corresponding to *P* < 1.0 × 10^-5^. Before applying this stringent multiple testing threshold, 80 cohort-specific protein associations, representing 52 unique proteins, passed all MR sensitivity analyses, with all 80 associations showing strong colocalization evidence with BMI-subtracted T2D (PP.H4 ≥ 0.8 by PWCoCo or SharePro; **Supplementary Table 10**).

Of these 52 proteins, 29 unique proteins, corresponding to 48 cohort-specific associations, additionally met the study-wide Bonferroni MR threshold of *P* <1 × 10^-5^ and were prioritized for downstream analyses (**Figure 2a**, **Table 1**, and **Supplementary Table 11**). Twelve proteins were supported by associations in at least two proteomic cohorts. NCAN replicated across all four cohorts; GCKR, HHIP, TBCE, TYRO3, and MANSC4 replicated across three cohorts. Six additional proteins, THG1L, SNUPN, RSPO3, PAM, LRIG1, and NUDT5, were supported in two cohorts. Effect directions were consistent across cohorts for all replicated proteins.

**Figure 2.**
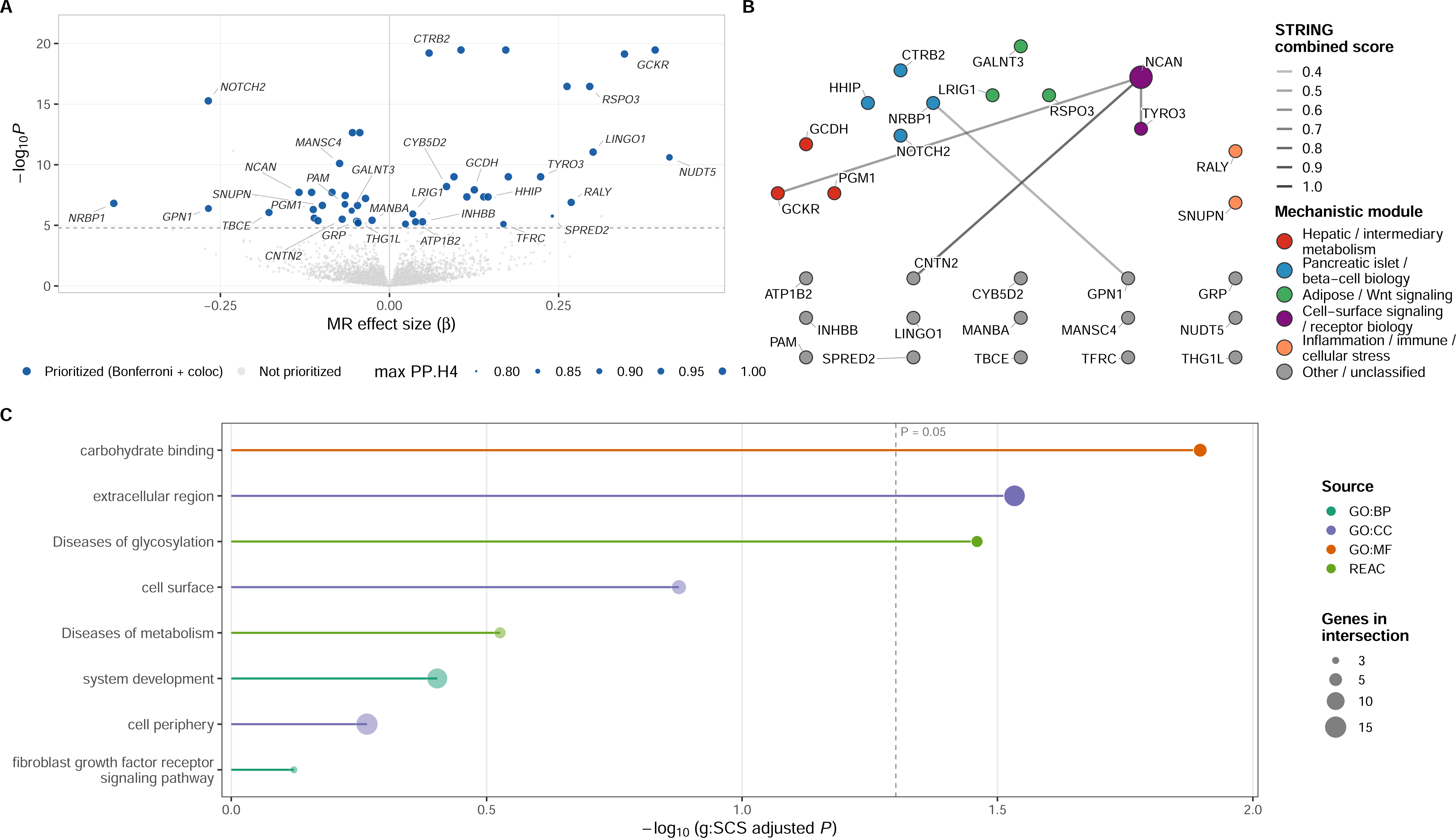
Putatively causal proteins for BMI-subtracted T2D and their mechanistic context. (A) Proteome-wide Mendelian randomization (MR) volcano plot. Each point is one MR test of a plasma protein on BMI-subtracted T2D in one cohort (ARIC, deCODE, Fenland, or UKB-PPP). The x-axis is the MR effect size (β, log-odds of BMI-subtracted T2D per SD higher protein) and the y-axis is -log_10_*P*-vaue. Navy blue points are the 29 prioritized proteins that pass study-wide Bonferroni significance (*P*< 0.05/4,991) and colocalized (PP.H4 > 0.8); grey points are all other tests. Point size is the maximum colocalization PP.H4 (PWCoCo or SharePro). The dashed line is the Bonferroni threshold; each prioritized protein is labelled once, at its most extreme effect-size across the four proteomics cohorts. (B) STRING protein-protein interaction network of the 29 prioritized proteins, with nodes coloured by mechanistic module and edges weighted by the STRING combined score. (C) Pathway enrichment of the 29 proteins (g:Profiler). For each ontology source (GO:BP, GO:CC, GO:MF, Reactome), up to the three most significant terms overlapping at least three of the prioritized proteins are shown; terms with no over-representation signal (g:SCS-adjusted *P* = 1) are omitted. Filled points denote the three terms passing the significance threshold (g:SCS-adjusted *P* < 0.05); the remaining terms are faded and shown for context. The x-axis represents -log_10_(g:SCS-adjusted *P*), dot size is the number of genes in the intersection, and the dashed line marks *P* = 0.05.

**Table 1.** Proteins associated with BMI-subtracted T2D that passed study-wide Bonferroni correction with strong colocalization evidence.

| Gene | # Cohorts | Cohort(s)* | MR $\beta$ (95% CI), $P^\dagger$ | PWCoCo PP.H4 | SharePro PP.H4 | Best eQTL tissue (PP.H4) | Druggability tier | Open Targets drug (max phase; indication)‡ |
| --- | --- | --- | --- | --- | --- | --- | --- | --- |
| GCKR | 3 | ARIC, deCODE, Fenland | $\beta_{\text{Fenland}} = 0.11$ (0.083 to 0.13); $P = 3.4 \times 10^{-20}$ | 1.00 | 1.00 | None $\geq 0.8$ in panel | Unclassified | — |
| CTRB2 | 1 | Fenland | $\beta_{\text{Fenland}} = 0.059$ (0.046 to 0.071); $P = 6.1 \times 10^{-20}$ | 1.00 | 1.00 | GTEx Pancreas (1.00); InsPIRE Islets (0.93) | Tier 3A | — |
| NUDT5 | 2 | Fenland, UKB-PPP | $\beta_{\text{Fenland}} = 0.35$ (0.27 to 0.42); $P = 7.4 \times 10^{-20}$ | 0.04 | 1.00 | METSIM Sub Adipose (0.97); GTEx Muscle Skeletal (0.94) | Unclassified | — |
| RSPO3 | 2 | ARIC, Fenland | $\beta_{\text{Fenland}} = 0.30$ (0.23 to 0.36); $P = 3.5 \times 10^{-17}$ | 0.98 | 1.00 | METSIM Sub Adipose (1.00); GTEx Adipose Sub (0.98) | Tier 3A | Rosmantuzumab (Phase 1; neoplasm) |
| NOTCH2 | 1 | UKB-PPP | $\beta_{\text{UKB-PPP}} = -0.27$ (-0.33 to -0.20); $P = 5.4 \times 10^{-16}$ | 0.96 | 1.00 | eQTLGen Whole Blood (0.96); GTEx Pancreas (0.93); GTEx Liver (0.92) | Tier 3A | Tarextumab (Phase 1; neoplasm incl. pancreatic carcinoma) |
| MANSC4 | 3 | ARIC, deCODE, Fenland | $\beta_{\text{Fenland}} = -0.055$ (-0.069 to -0.040); $P = 2.2 \times 10^{-13}$ | 0.57 | 1.00 | None $\geq 0.8$ in panel | Unclassified | — |
| LINGO1 | 1 | deCODE | $\beta_{\text{deCODE}} = 0.30$ (0.21 to 0.39); $P = 9.1 \times 10^{-12}$ | 0.03 | 1.00 | None $\geq 0.8$ in panel | Tier 1 | Opicinumab (Phase 2; multiple sclerosis) |
| TYRO3 | 3 | ARIC, deCODE, Fenland | $\beta_{\text{deCODE}} = 0.22$ (0.15 to 0.30); $P = 9.9 \times 10^{-10}$ | 0.80 | 1.00 | None $\geq 0.8$ in panel | Tier 1 | — |
| CYB5D2 | 1 | ARIC | $\beta_{\text{ARIC}} = 0.084$ (0.056 to 0.11); $P = 6.3 \times 10^{-9}$ | 0.01 | 1.00 | METSIM Sub Adipose (0.95); InsPIRE Islets (0.80) | Tier 3B | — |
| GCDH | 1 | ARIC | $\beta_{\text{ARIC}} = 0.13$ (0.082 to 0.17); $P = 1.2 \times 10^{-8}$ | 0.93 | 1.00 | GTEx Liver (0.99); GTEx Pancreas (0.99); eQTLGen Whole Blood (0.98) | Unclassified | — |
| NCAN | 4 | ARIC, deCODE, Fenland, UKB-PPP | $\beta_{\text{ARIC}} = -0.085$ (-0.11 to -0.055); $P = 1.9 \times 10^{-8}$ | 0.02 | 1.00 | None $\geq 0.8$ in panel | Tier 3A | — |
| SNUPN | 2 | ARIC, Fenland | $\beta_{\text{ARIC}} = -0.065$ (-0.089 to -0.042); $P = 3.5 \times 10^{-8}$ | 0.98 | 1.00 | eQTLGen Whole Blood (0.99); GTEx Adipose Visceral Omentum (0.97); GTEx Whole Blood (0.96) | Unclassified | — |
| HHIP | 3 | ARIC, deCODE, Fenland | $\beta_{\text{deCODE}} = 0.15$ (0.094 to 0.20); $P = 4.5 \times 10^{-8}$ | 1.00 | 1.00 | None $\geq 0.8$ in panel | Tier 3A | — |
| PAM | 2 | ARIC, deCODE | $\beta_{\text{ARIC}} = -0.035$ (-0.048 to -0.023); $P = 6.0 \times 10^{-8}$ | 1.00 | 1.00 | GTEx Whole Blood (0.96) | Tier 2 | — |
| RALY | 1 | UKB-PPP | $\beta_{\text{UKB-PPP}} = 0.27$ (0.17 to 0.37); $P = 1.3 \times 10^{-7}$ | 0.96 | 0.99 | None $\geq 0.8$ in panel | Unclassified | — |
| NRBP1 | 1 | deCODE | $\beta_{\text{deCODE}} = -0.41$ (-0.56 to -0.26); $P = 1.5 \times 10^{-7}$ | 0.01 | 1.00 | None $\geq 0.8$ in panel | Tier 3B | — |
| TBCE | 3 | ARIC, deCODE, Fenland | $\beta_{\text{ARIC}} = -0.047$ ($-0.065$ to $-0.029$); $P = 2.3 \times 10^{-7}$ | 0.97 | 0.99 | None $\geq 0.8$ in panel | Unclassified | — |
| GPN1 | 1 | deCODE | $\beta_{\text{deCODE}} = -0.27$ ($-0.37$ to $-0.16$); $P = 4.1 \times 10^{-7}$ | 0.01 | 0.96 | GTEx Muscle Skeletal (0.95) | Unclassified | — |
| PGM1 | 1 | Fenland | $\beta_{\text{Fenland}} = -0.11$ ($-0.16$ to $-0.069$); $P = 5.0 \times 10^{-7}$ | 0.99 | 0.98 | None $\geq 0.8$ in panel | Unclassified | — |
| GALNT3 | 1 | UKB-PPP | $\beta_{\text{UKB-PPP}} = -0.056$ ($-0.078$ to $-0.034$); $P = 6.1 \times 10^{-7}$ | 0.91 | 0.91 | eQTLGen Whole Blood (0.99); GTEx Adipose Sub (0.98); METSIM Sub Adipose (0.96) | Unclassified | — |
| LRIG1 | 2 | ARIC, Fenland | $\beta_{\text{Fenland}} = 0.034$ ($0.021$ to $0.048$); $P = 1.1 \times 10^{-6}$ | 0.97 | 0.90 | METSIM Sub Adipose (0.96) | Unclassified | — |
| SPRED2 | 1 | UKB-PPP | $\beta_{\text{UKB-PPP}} = 0.24$ ($0.14$ to $0.34$); $P = 1.7 \times 10^{-6}$ | 0.02 | 0.81 | None $\geq 0.8$ in panel | Unclassified | — |
| CNTN2 | 1 | Fenland | $\beta_{\text{Fenland}} = -0.070$ ($-0.099$ to $-0.041$); $P = 3.1 \times 10^{-6}$ | 0.93 | 0.99 | None $\geq 0.8$ in panel | Tier 3B | — |
| MANBA | 1 | ARIC | $\beta_{\text{ARIC}} = -0.026$ ($-0.037$ to $-0.015$); $P = 3.8 \times 10^{-6}$ | 0.98 | 1.00 | METSIM Sub Adipose (1.00); InsPIRE Islets (0.99); eQTLGen Whole Blood (0.87) | Tier 2 | — |
| GRP | 1 | UKB-PPP | $\beta_{\text{UKB-PPP}} = -0.049$ ($-0.070$ to $-0.028$); $P = 4.4 \times 10^{-6}$ | 0.97 | 0.91 | None $\geq 0.8$ in panel | Tier 3B | — |
| THG1L | 2 | deCODE, Fenland | $\beta_{\text{Fenland}} = -0.046$ ($-0.066$ to $-0.027$); $P = 4.9 \times 10^{-6}$ | 0.98 | 0.93 | None $\geq 0.8$ in panel | Unclassified | — |
| INHBB | 1 | deCODE | $\beta_{\text{deCODE}} = 0.049$ ($0.028$ to $0.070$); $P = 5.1 \times 10^{-6}$ | 1.00 | 0.00 | None $\geq 0.8$ in panel | Tier 3A | — |
| ATP1B2 | 1 | ARIC | $\beta_{\text{ARIC}} = 0.039$ ($0.022$ to $0.055$); $P = 5.1 \times 10^{-6}$ | 0.98 | 0.93 | GTEx Adipose Sub (0.96); GTEx Muscle Skeletal (0.95); InsPIRE Islets (0.94) | Tier 3B | Istaroxime (Phase 2; heart failure) |
| TFRC | 1 | ARIC | $\beta_{\text{ARIC}} = 0.17$ ($0.095$ to $0.24$); $P = 7.9 \times 10^{-6}$ | 0.92 | 0.93 | None $\geq 0.8$ in panel | Tier 1 | Pabinafusp alfa (Phase 3; mucopolysaccharidosis II) |

One representative row per protein, ordered by ascending $P$ . # Cohorts is the number of proteomic cohorts in which the protein passed the MR Bonferroni threshold ( $P < 0.05/4,991$ ) and met the *cis*-pQTL colocalization criterion (PP.H4 $\geq 0.8$ by PWCoCo or SharePro). PWCoCo and SharePro PP.H4 are the maxima across the listed cohorts. Best eQTL tissue lists up to the top three tissues showing *cis*-eQTL-*cis*-pQTL colocalization (PP.H4 $\geq 0.8$ ) across GTEx, InsPIRE, METSIM, and eQTLGen. Druggability tier follows the classification of Finan *et al.*; proteins absent from that source are listed as Unclassified.
\*Proteomic cohorts in which the *cis*-pQTL instrument and the downstream MR and colocalization criteria were both satisfied. † MR $\beta$ (95% CI) and $P$ are from the cohort with the smallest $P$ (indicated in subscript); cohort-specific estimates for all cohorts are provided in **Supplementary Table 1**. ‡ Highest-phase drug in Open Targets targeting the protein, with primary indication(s); "—" indicates no clinical-stage drug at the time of analysis.
BMI, body mass index; CI, confidence interval; *cis*-eQTL, *cis* expression quantitative trait locus; *cis*-pQTL, *cis* protein quantitative trait locus; MR, Mendelian randomization; PP.H4, posterior probability of a shared causal variant; PWCoCo, pairwise conditional and colocalization analysis; T2D, type 2 diabetes.

Several prioritized proteins mapped to established pathways in glucose metabolism and diabetes susceptibility. GCKR, encoding the glucokinase regulatory protein, was strongly associated with higher BMI-subtracted T2D liability across ARIC, deCODE, and Fenland, with consistent effect directions and strong colocalization evidence across cohorts (OR_deCODE_ 1.48, 95% CI 1.36–1.61; *P* = 3.4 × 10^-20^; PP.H4 = 1.00). NOTCH2, a regulator of pancreatic β-cell development and function, showed an inverse association in UKB-PPP (OR 0.77, 95% CI 0.72–0.82; *P* = 5.4 × 10^-16^; PP.H4 = 0.96). HHIP was associated with higher BMI-subtracted T2D liability across ARIC, deCODE, and Fenland (OR_deCODE_ = 1.16, 95% CI 1.10–1.22; *P* = 4.5 × 10^-8^; PP.H4 = 1.00), while CTRB2 showed a strong association in Fenland (OR_Fenland_ = 1.06, 95% CI 1.05–1.07; *P* = 6.1 × 10^-20^; PP.H4 = 1.00), consistent with pancreatic biology. Additional signals highlighted broader metabolic and signaling pathways such as PGM1 and GCDH related to hepatic intermediary metabolism, RSPO3 and GALNT3 related to adipose or Wnt signaling, and TYRO3 and LRIG1 representing receptor-mediated signaling pathways.

### Functional Annotation of Candidate Proteins

To aid biological interpretation, we grouped the 29 prioritized proteins into literature-supported categories reflecting major axes of T2D pathophysiology (**Supplementary Table 12**). These included hepatic and intermediary metabolism (GCKR, PGM1, GCDH), pancreatic islet and β-cell biology (NOTCH2, CTRB2, HHIP, NRBP1), adipose tissue function and Wnt signaling (RSPO3, GALNT3, LRIG1), cell-surface signaling and receptor biology (TYRO3, NCAN), and inflammation, immune regulation, and cellular stress (SNUPN, RALY). The remaining 15 proteins had more heterogeneous enzymatic, transport, or stress-response annotations and were classified as other or unclassified in the mechanistic map.

STRING v12 analysis showed limited protein-protein connectivity among the prioritized proteins. Using medium-confidence interactions, we observed four physical or functional interactions among the 29 proteins from NCAN–TYRO3, NCAN–GCKR, NCAN–CNTN2, and GPN1–NRBP1 (**Figure 2b** and **Supplementary Table 13**). Six proteins contributed at least one interaction edge, and the whole-network protein–protein interaction enrichment test was not significant (*P* = 0.19). These findings suggest that the prioritized proteins do not represent a single tightly connected molecular complex but instead reflect multiple biological processes contributing to BMI-subtracted T2D susceptibility.

Pathway over-representation analysis using the annotated genome as the background identified three enriched terms after multiple-testing correction (**Figure 2c** and **Supplementary Table 14**) pertaining to a Gene Ontology “extracellular region” (14 of 27 mapped genes; adjusted *P* = 0.029), Gene Ontology “carbohydrate binding” (5 of 27; adjusted *P* = 0.013), and Reactome “Diseases of glycosylation” (4 of 27; adjusted *P* = 0.035). The carbohydrate-binding and glycosylation-related terms were driven by GALNT3, NCAN, NOTCH2, and PGM1, supporting a potential contribution of glycosylation-related biology to the prioritized protein set.

### eQTL Colocalization Supports Regulatory Evidence in Diabetes-Relevant Tissues

We performed tissue-specific *cis*-eQTL colocalization analyses across GTEx, InsPIRE, METSIM, and eQTLGen to identify candidate tissues of action relevant to T2D biology (**Figure 3** and **Supplementary Table 15**). Overall, 13 of the 29 prioritized proteins showed strong eQTL colocalization evidence (PP.H4 ≥ 0.8) in at least one tissue.

**Figure 3.**
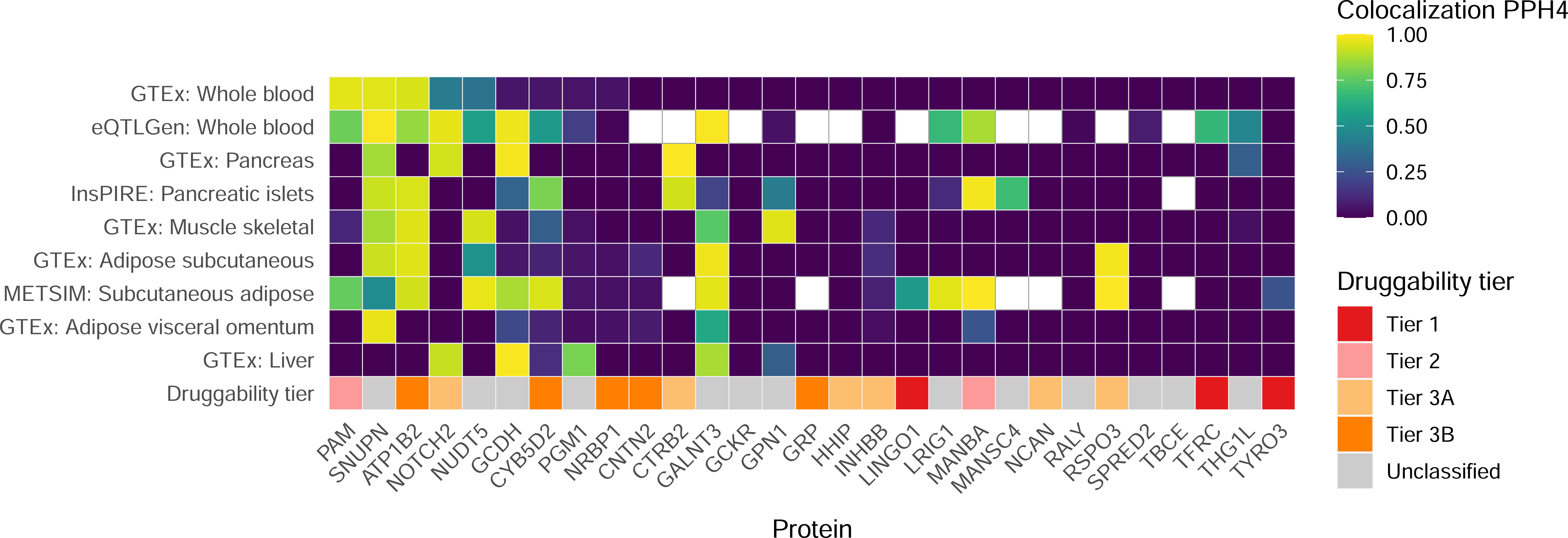
Colocalization and druggability heatmap of MR-prioritized proteins. Heatmap of pQTL and eQTL colocalization posterior probability (PP.H4) for each candidate protein (columns) across nine expression datasets (rows): six T2D-relevant GTEx tissues (whole blood, pancreas, skeletal muscle, subcutaneous adipose, visceral adipose [omentum], and liver) plus three tissue-specialty eQTL sources (InsPIRE pancreatic islets, METSIM subcutaneous adipose, and eQTLGen whole blood). Tile colour is the colocalization PP.H4 (viridis scale); white tiles with a grey border indicate genes not measured in those resources and are therefore not assessed for colocalization. Dark tiles are those that were tested for colocalization but did not colocalize (PP.H4 = 0). The bottom annotation row is the druggability row which uses fixed colors: Tier 1, red; Tier 2, pink; Tier 3A, light orange; Tier 3B, orange; Unclassified, grey.

Several proteins implicated hepatic and intermediary metabolic pathways. GCDH showed strong colocalization in liver (GTEx Liver PP.H4 = 0.99), consistent with its role in hepatic intermediary metabolism, and also colocalized in pancreas (PP.H4 = 0.99) and eQTLGen whole blood (PP.H4 = 0.98). NOTCH2 showed broader tissue colocalization, including pancreas (PP.H4 = 0.93), liver (PP.H4 = 0.92), and eQTLGen whole blood (PP.H4 = 0.96), supporting regulatory evidence across multiple tissues relevant to T2D pathophysiology. By contrast, GCKR and PGM1, despite their biological links to hepatic glucose metabolism, did not colocalize in the evaluated tissue panel.

Pancreatic and islet-related regulatory signals were also observed. CTRB2 colocalized with both GTEx pancreas (PP.H4 = 1.00) and InsPIRE pancreatic islets (PP.H4 = 0.93), supporting a pancreas-specific regulatory mechanism. MANBA colocalized with InsPIRE pancreatic islets (PP.H4 = 0.99), and ATP1B2 and CYB5D2 showed additional islet colocalization (InsPIRE PP.H4 = 0.94 and 0.80, respectively). SNUPN also colocalized in InsPIRE islets (PP.H4 = 0.92) as part of a broader multi-tissue signal.

Adipose-related regulatory evidence was observed for several proteins. RSPO3, a regulator of adipose Wnt signaling, showed strong colocalization in both METSIM subcutaneous adipose tissue (PP.H4 = 1.00) and GTEx subcutaneous adipose tissue (PP.H4 = 0.98). MANBA showed its strongest signal in METSIM subcutaneous adipose tissue (PP.H4 = 1.00), in addition to its pancreatic-islet and whole-blood signals. GALNT3 showed broader colocalization across liver (PP.H4 = 0.87), subcutaneous adipose tissue (GTEx PP.H4 = 0.98; METSIM PP.H4 = 0.96), and eQTLGen whole blood (PP.H4 = 0.99). CYB5D2 also colocalized in METSIM subcutaneous adipose tissue (PP.H4 = 0.95). LRIG1 colocalized in METSIM subcutaneous adipose tissue (PP.H4 = 0.96). NUDT5 colocalized in both METSIM subcutaneous adipose tissue (PP.H4 = 0.97) and GTEx skeletal muscle (PP.H4 = 0.94).

Whole blood regulatory signals were identified for several proteins. PAM (GTEx whole blood PP.H4 = 0.96), SNUPN (eQTLGen PP.H4 = 0.99; GTEx whole blood PP.H4 = 0.96), ATP1B2 (GTEx whole blood PP.H4 = 0.94; eQTLGen PP.H4 = 0.84), and MANBA (eQTLGen PP.H4 = 0.87) colocalized in whole-blood resources, suggesting that circulating-cell regulatory mechanisms may contribute to a subset of BMI-subtracted T2D signals. SNUPN showed the broadest cross-tissue regulatory pattern, colocalizing in seven tissues, including pancreas (PP.H4 = 0.87), pancreatic islets (InsPIRE PP.H4 = 0.91), skeletal muscle (PP.H4 = 0.87), subcutaneous and visceral adipose tissue (PP.H4 = 0.92 and 0.97), and both whole-blood resources.

Notably, 16 of the 29 prioritized proteins, including GCKR, HHIP, NCAN, TBCE, and GRP, did not colocalize with any *cis*-eQTL in the nine evaluated tissue resources. Additional proteins without *cis*-eQTL colocalization included CNTN2, INHBB, LINGO1, MANSC4, NRBP1, PGM1, RALY, SPRED2, TFRC, THG1L, and TYRO3. This may reflect incomplete tissue coverage in currently available eQTL datasets, post-transcriptional regulation of circulating protein abundance, or limited statistical power in smaller tissue-specific resources.

### Actionability assessment of target proteins

We assessed therapeutic actionability for the 29 prioritized proteins using the druggable genome and the Open Targets Platform. Sixteen of the 29 proteins (55%) were classified within the druggable genome with three in Tier 1 (LINGO1, TYRO3, TFRC; approved or clinical-stage targets), two in Tier 2 (PAM, MANBA; closely related to known drug targets), six in Tier 3A (CTRB2, RSPO3, NOTCH2, NCAN, HHIP, INHBB; secreted or extracellular proteins with biotherapeutic potential), and five in Tier 3B (CYB5D2, NRBP1, CNTN2, GRP, ATP1B2; other extracellular proteins); the remaining 13 proteins were unclassified (**Figure 3**).

Open Targets queries identified existing compounds against 5 of the 29 prioritized proteins (TFRC, ATP1B2, LINGO1, NOTCH2, RSPO3), yielding 27 protein-drug pairs after excluding compounds that had already reached completed Phase 3 or Phase 4 trials (**Table 1, Supplementary Table 16** and **17**). Among these, RSPO3 and NOTCH2 represented examples in which existing pharmacology aligned with MR-prioritized biology. Rosmantuzumab, an RSPO3–LGR4 inhibitor evaluated in early-phase oncology trials, mapped to RSPO3, for which genetically higher circulating protein levels were associated with increased BMI-subtracted T2D liability and showed strong colocalization in subcutaneous adipose tissue (METSIM PP.H4 = 1.00; GTEx PP.H4 = 0.98). Tarextumab, a NOTCH2-targeting antibody developed in oncology, mapped to NOTCH2, for which genetically lower circulating protein levels were associated with lower BMI-subtracted T2D liability. Other prioritized Tier 1 proteins, including TYRO3, LINGO1, and TFRC, were linked to clinical-stage compounds developed for non-metabolic indications.

## Conclusions

In this study, we used GWAS-by-subtraction to partition type 2 diabetes (T2D) genetic liability into BMI-related and BMI-subtracted components, then applied proteome-wide MR across four independent plasma proteomic cohorts to identify circulating proteins putatively causal for BMI-subtracted T2D. This approach prioritized 29 candidate proteins. Tissue-resolution *cis*-eQTL colocalization across GTEx, pancreatic islets, subcutaneous adipose, and whole blood implicated hepatic, pancreatic-islet, adipose, and immune-related compartments as plausible tissues of action for several targets. Actionability analyses further showed that 16 of the 29 prioritized proteins were classified within the druggable genome, and Open Targets identified existing clinical-stage compounds engaging five proteins. Together, these findings provide a genetically informed shortlist of candidate targets that may inform therapeutic development or repurposing strategies for T2D pathways not fully explained by adiposity.

These findings matter clinically because BMI does not capture all biological pathways leading to T2D. Although GLP-1 receptor agonists, dual incretin agonists, and SGLT2 inhibitors have reshaped diabetes treatment, some patients have inadequate response, intolerance, contraindications, access barriers, or develop T2D without obesity (2,3,36). Protein targets identified through BMI-subtracted T2D genetics may therefore point to mechanisms that complement existing therapies, particularly for subgroups in which BMI-linked pathways are less central.

The BMI-subtracted T2D component represents genetic liability to T2D that is not fully captured by BMI. This is clinically relevant because T2D risk and treatment response vary among individuals with similar BMI, and because many patients develop T2D without obesity or do not achieve adequate glycemic control with weight-centered therapies. By prioritizing proteins associated with this component, our analysis aimed to identify biological pathways that may complement adiposity-targeted approaches. The tissue-specific regulatory evidence further suggests that the BMI-subtracted component reflects metabolic biology beyond BMI, with relevance to insulin secretion, hepatic metabolism, adipose function, and inflammation.

Several prioritized proteins mapped to pathways with established relevance to T2D. GCKR, which encodes glucokinase regulatory protein, modulates hepatic glucose and lipid metabolism by inhibiting glucokinase (37); here, genetically higher circulating GCKR was associated with higher BMI-subtracted T2D liability across three cohorts. The absence of detectable GCKR eQTL colocalization may reflect post-transcriptional regulation of circulating protein levels, limited power in available liver eQTL resources, or regulatory contexts. NOTCH2, a regulator of pancreatic development and β-cell biology (38), showed a protective genetic association in UKB-PPP and colocalized with *cis*-eQTLs in pancreas, liver, and whole blood. CTRB2, linked to pancreatic exocrine biology and islet pathology, showed strong pancreatic and islet colocalization, while RSPO3, an adipose-related Wnt signaling regulator (39), colocalized in subcutaneous adipose tissue.

The broader protein set suggested mechanisms extending beyond individual established T2D genes. Pathway over-representation analyses highlighted carbohydrate binding and Reactome diseases of glycosylation, driven by GALNT3, NCAN, NOTCH2, and PGM1. Notably, the prioritized proteins did not form a densely connected protein-protein interaction network, suggesting that BMI-subtracted T2D liability reflects multiple partially distinct mechanisms rather than a single molecular complex. Whether the glycosylation-related findings represent a shared mechanistic axis or several independent pathways converging on a common ontology will require targeted experimental evaluation.

The actionability analysis provides hypotheses for therapeutic development, but it also demonstrates the need for careful direction-of-effect interpretation. RSPO3 provides the clearest mechanism-aligned repurposing example among the targets with existing clinical-stage pharmacology. Genetically higher circulating RSPO3 was associated with higher BMI-subtracted T2D liability and showed strong colocalization in adipose tissue. Because rosmantuzumab blocks the RSPO3–LGR4 interaction, its pharmacologic direction is consistent with the protective direction predicted by the genetic evidence (40).

Several limitations should be considered. First, the analyses were restricted to European ancestry because currently available non-European ancestry T2D GWAS have limited sample sizes and statistical power. The prioritized proteins should be evaluated in ancestrally diverse datasets to assess generalizability. Second, GWAS-by-subtraction removes genetic associations shared with BMI but cannot eliminate all adiposity-related biology. Residual effects related to waist-to-hip ratio, ectopic fat, fat distribution, or other traits may persist. Third, plasma *cis*-pQTL MR captures genetically predicted circulating protein abundance, which may not reflect tissue-specific protein activity or intracellular mechanisms. Fourth, the eQTL resources used here do not capture the full range of tissue, cellular, and disease-state contexts relevant to T2D. This may explain why several strongly prioritized proteins lacked eQTL colocalization despite MR and pQTL colocalization evidence. Finally, the actionability assessment identifies existing pharmacology but does not establish that the drug mechanism, dose, tissue exposure, or direction of target modulation will reproduce the genetically predicted effect. Functional studies in T2D-relevant tissues and careful pharmacologic validation will be required.

In conclusion, by separating BMI-shared from BMI-subtracted components of T2D genetic liability and integrating proteome-wide MR with colocalization, we identified 29 circulating proteins associated with T2D risk not fully explained by BMI. These proteins implicate hepatic, pancreatic-islet, adipose, immune, and glycosylation-related pathways, and several are already connected to clinical-stage pharmacology. These findings nominate genetically supported protein targets for follow-up studies of T2D therapies that act through mechanisms beyond weight loss.

## Declarations

## Acknowledgments

C.Y.-S. acknowledges the Social Sciences Computing Core at the University of Wisconsin-Madison for providing computing resources for conducting these analyses.

## Author Contributions

C.-Y.S. and T.L. conceived and designed the study and interpreted the results. C.-Y.S. performed the analyses and drafted the manuscript. C.-Y.S. and T.L. critically revised the manuscript for important intellectual content. All authors approved the final version of the manuscript. C.-Y.S. is the guarantor of this work.

## Prior Presentation

This study has not been presented previously.

