## Supplementary Note 1 for "Disentangling adiposity-related and non-adiposity-related genetic pathways for type 2 diabetes"

**Supplementary Note 1. Data sources and cohort characteristics.**

| **Resource** | **Trait or measurement** | **Platform** | **Sample size (European ancestry)** |
| --- | --- | --- | --- |
| T2D GWAS (1) | Type 2 diabetes (cases vs controls; unadjusted for BMI) | Imputed genotypes meta-analysis | 74,124 cases / 824,006 controls (n = 898,130) |
| BMI GWAS (2) | Body mass index | GIANT + UK Biobank meta-analysis | n = 681,275 |
| ARIC (3) | Plasma proteomic levels (*cis*-pQTL) | SomaScan v4 (~5,000 proteins) | n = 7,213; 4,657 proteins |
| deCODE (4) | Plasma proteomic levels (*cis*-pQTL) | SomaScan v4 (~5,000 proteins) | n = 35,559; 4,719 proteins |
| Fenland (5) | Plasma proteomic levels (*cis*-pQTL) | SomaScan v4 (~5,000 proteins) | n = 10,708; 4,775 proteins |
| UK Biobank PPP (6) | Plasma proteomic levels (*cis*-pQTL) | Olink Explore 3072 (~2,900 proteins) | n = 34,557; 2,923 proteins |
| GTEx v8 (7) | *cis*-eQTL in 6 T2D-relevant tissues | RNA-seq | Multi-tissue; tissue sample sizes per (16) |
| InsPIRE (8) | *cis*-eQTL in pancreatic islets | RNA-seq | n = 420 donors |
| METSIM (9) | *cis*-eQTL in subcutaneous adipose | RNA-seq | n = 434 Finnish men |
| eQTLGen (Phase I) (10) | *cis*-eQTL in whole blood | Expression arrays / RNA-seq meta-analysis | up to n = 31,684 |

BMI, body mass index; eQTL, expression quantitative trait locus; GWAS, genome-wide association study; PPP, Pharma Proteomics Project; pQTL, protein quantitative trait locus; T2D, type 2 diabetes.
